# DEVELOPMENT AND INTERNAL EVALUATION OF MODELS FOR PREDICTING PROLONGED HOSPITALISATION AMONG PATIENTS WITH SICKLE CELL DISEASE IN A PUBLIC UNIVERSITY HOSPITAL IN GHANA

**DOI:** 10.64898/2026.11.10.26367799

**Authors:** Jerry Selase Sifa, Gideon Yormabu, Gifty Hariet Odame, Jessie Amoako Quayson, Wendy Asamoah Boadu, Mark Hammond, Nina Nerkai Ashong, Afua Asabea Amoabeng Nti

## Abstract

**Background:** Prolonged hospitalisation is an important burden among patients with sickle cell disease (SCD), with implications for healthcare utilisation and patient and caregiver burden. Early prediction may support timely clinical and resource-management interventions.

**Objective:** To identify factors associated with prolonged hospitalisation among patients with SCD and compare the predictive performance of multivariable logistic regression with five supervised machine-learning algorithms.

**Methods:** This retrospective study included SCD inpatients aged ≥16 years admitted to the University of Ghana Hospital between January 2019 and December 2024. Logistic regression, random forest, XGBoost, support vector machine, and k-nearest neighbours models were developed using five-fold stratified cross-validation. Class weighting and SMOTE were applied within training folds. Model performance was evaluated using AUC-ROC, AUC-PR, sensitivity, specificity, positive predictive value (PPV), negative predictive value (NPV), F1 score, and balanced accuracy.

**Results:** Among 872 patients, 137 (15.7%) experienced prolonged hospitalisation. Acute chest syndrome (aOR=4.28; 95% CI: 2.31–7.94), malaria (aOR=3.77; 95% CI: 2.11–6.74), pneumonia (aOR=3.45; 95% CI: 2.02–5.90), and anaemia (aOR=2.71; 95% CI: 1.13–6.51) were independently associated with prolonged hospitalisation, whereas higher haemoglobin was protective (aOR=0.86; 95% CI: 0.78–0.96). Logistic regression achieved the highest AUC-ROC (0.692), F1 score (0.418) and balanced accuracy (0.677), while random forest achieved the highest AUC-PR (0.375). KNN had the highest sensitivity (59.3%) and XGBoost the highest specificity (87.2%). AUC-ROC confidence intervals overlapped substantially across models.

**Conclusion:** Infection-related complications and anaemia were important predictors of prolonged hospitalisation, while higher haemoglobin was protective. ML models showed modest performance without clear superiority over logistic regression. Larger multicentre studies with external validation are warranted.

## INTRODUCTION

Sickle cell disease (SCD) remains a major global health challenge as affected individuals experience a lifelong, severely disabling condition associated with reduced quality of life, high healthcare utilization, substantial economic burden and premature mortality(1,2). In 2021, an estimated 7.74 million people were living with SCD of which 515,000 were babies(2). Sub-Saharan Africa accounts for nearly 80% of sickle cell disease births. In the same year, SCD was responsible for approximately 376,000 deaths of which 81,100 occurred among children, making it the 12th leading cause of mortality globally(3). It is the leading cause of death among children and adolescents in Sub-Saharan Africa(3).

Length of hospital stay (LOS) is a widely used proxy for disease severity, complication burden and healthcare resource utilisation(4). Prolonged hospitalisation is associated with increased healthcare costs, greater risk of nosocomial complications, and substantial burden on patients and caregivers, making early identification of patients at risk of prolonged stay clinically and operationally valuable(5,6).

A recently published study in Ghana reported that 16% of SCD inpatients spent over 7 days in the hospital(7). Multivariate logistic regression identified comorbidities, infection, acute chest syndrome, hyperhemolysis, sequestration crisis, and the need for transfusion as predictors of prolonged hospital stay among SCD inpatients(7).

Traditional regression approaches like logistic regression, while interpretable, assume predominantly linear and additive relationships between predictors and outcome(8). These may not capture higher-order interactions among clinical and haematological variables. Supervised machine learning (ML) approaches offer potentially greater flexibility in modelling such relationships, though this must be weighed against reduced interpretability and, at modest sample sizes, a risk of overfitting(9).

This study therefore seeks to assess to identify the factors accounting for prolonged hospitalisation (>7 days) among SCD inpatients using multivariable logistic regression, and compare the predictive performance of five supervised ML algorithms against conventional logistic regression for this outcome.

## METHODS AND MATERIALS

### Study Design and Setting

This retrospective study analysed the medical records of patients diagnosed with sickle cell disease who were admitted to the University of Ghana Hospital. Records of all SCD inpatients admitted between January 2019 and December 2024 were extracted from the Hospital Information System (HIS). The extracted variables included demographic characteristics (age at admission and sex) and clinical characteristics, including acute chest syndrome, anaemia, malaria, pneumonia, and haemoglobin level. Length of hospital stay was the primary outcome and was used to classify patients into non-prolonged (≤7 days) and prolonged hospitalisation (>7 days).

### Study Population

Patients with a confirmed diagnosis of sickle cell disease (SCD) who were admitted as inpatients during the study period were eligible for inclusion. The inclusion criteria were being aged 16 years or older and having documented admission and discharge dates from which length of hospital stay (LOS) could be determined. Records of patients younger than 16 years was excluded. Records with missing information on the outcome variable were also excluded. Following these exclusions, 872 patients with complete data on all variables included in the analysis constituted the final analytic sample.

**Figure 1.**
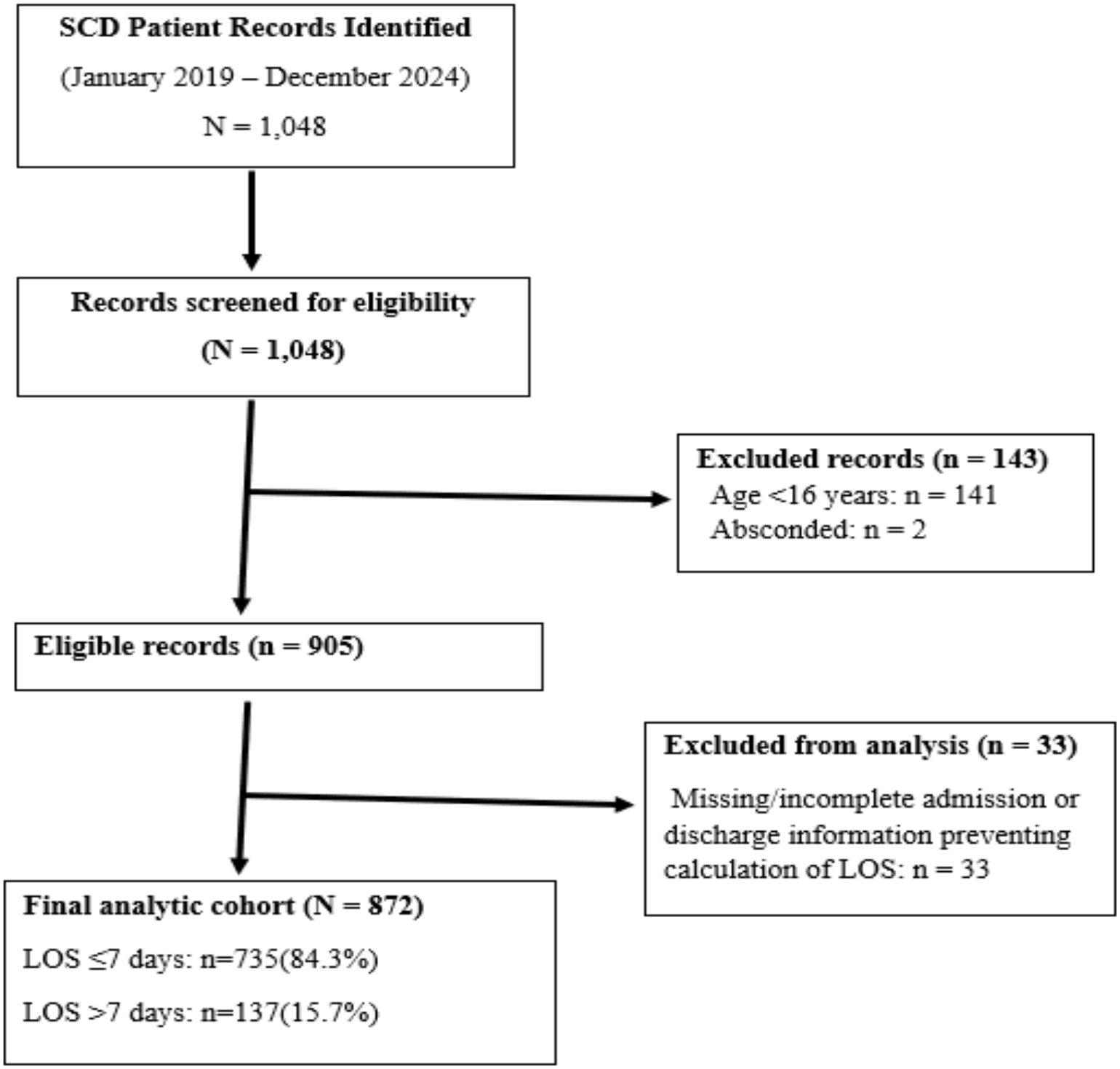
Flow diagram of SCD patient record selection and inclusion in the analytic cohort.

### Outcome Definition

The primary outcome was prolonged hospitalisation, defined a priori as a length of stay (LOS) exceeding 7 days(7). Length of stay was defined as the number of days between the documented date of admission and date of discharge. Prediction was performed using clinical information available within the first 48 hours of admission.

### Predictor Variables

Candidate predictors comprised acute chest syndrome, anaemia, malaria, pneumonia, haemoglobin (Hb) and age at admission. The variables were required to be supported by literature and available within the first 48 hours of admission to ensure that the model could theoretically be used for early risk stratification. All variables were extracted from inpatient clinical records. Binary variables were coded (present=1; absent=0). Haemoglobin and age were retained as continuous variables. The average VIF was 1.13 (min=1.00; max=1.09).

### Data Preprocessing

Given the observed outcome prevalence of 15.7%, class imbalance was addressed using class weighting and the Synthetic Minority Oversampling Technique (SMOTE). These techniques were applied exclusively within training folds during cross-validation to avoid data leakage into validation/test partitions. Continuous predictors were standardised (zero mean, unit variance) prior to fitting distance-based algorithms (support vector machine, k-nearest neighbours).

### Model Development

Five supervised classification algorithms were developed and compared: logistic regression (also serving as the interpretable clinical benchmark), random forest, extreme gradient boosting (XGBoost), support vector machine (SVM) and k-nearest neighbours (KNN). The models were developed and tuned using five-fold stratified cross-validation within the training set (80%) and subsequently evaluated on an independent held-out test set (20%). Model hyperparameters were optimized using the grid search method.

### Model Evaluation

Model discrimination was evaluated using the area under the receiver operating characteristic curve (AUC-ROC) and the area under the precision-recall curve (AUC-PR). The AUC-PR curve because of the class imbalance in the outcome. Additional metrics reported include sensitivity, specificity, positive predictive value (PPV), negative predictive value (NPV), F1-score and balanced accuracy. Point estimates and 95% confidence intervals for key metrics were derived via bootstrap resampling (1,000 resamples) of the test-set predictions.

### Model Interpretability

Permutation feature importance was computed for each algorithm to quantify the contribution of individual predictors to model discrimination (AUC-ROC) enabling comparison of predictor rankings across algorithms on a common scale.

### Data Management

Data were exported in CSV format and cleaned and coded using Microsoft Excel (version 2019). The cleaned dataset was imported into Stata 14 *(StataCorp LLC, College Station, TX, USA)* for statistical analysis. Bivariate analyses were performed to examine differences in characteristics between patients with and without prolonged hospitalisation. The independent-samples *t* test was used to compare continuous variables, while Pearson’s chi-square test or Fisher’s exact test was used to compare categorical variables, as appropriate. The events-per-variable (EPV) ratio was approximately 23 (137 prolonged-hospitalisation events/6 candidate predictors)

Machine learning model development, tuning and evaluation were conducted in Python [version 3.13] using scikit-learn, XGBoost, and imbalanced-learn packages in Google Colab.

### Ethical Considerations

This study utilized de-identified routine care data. Ethical approval was obtained from the Noguchi Memorial Institute for Medical Research Institutional Review Board (NMIMR-IRB) (Study No. 111/25-26). The requirement for informed consent was waived due to the retrospective nature of the study and the use of de-identified data.

This study is reported in accordance with the TRIPOD+AI guideline for studies developing or validating multivariable prediction models.

## RESULTS

### Baseline Characteristics

A total of 872 SCD inpatients with complete data were included in the analysis. Overall, 137 patients (15.7%; 95% CI: 13.4% to 18.2%) experienced prolonged hospitalization [Table 1]. The mean age was 26.1 years (SD = 8.7), and 53.0% (n = 458) of the patients were female. Bivariate analysis showed significant associations between prolonged hospitalization and following clinical factors: acute chest syndrome (p < 0.001), anaemia (p < 0.001), malaria (p < 0.001), pneumonia (p < 0.001), and haemoglobin level (p < 0.001).

**Table 1.**
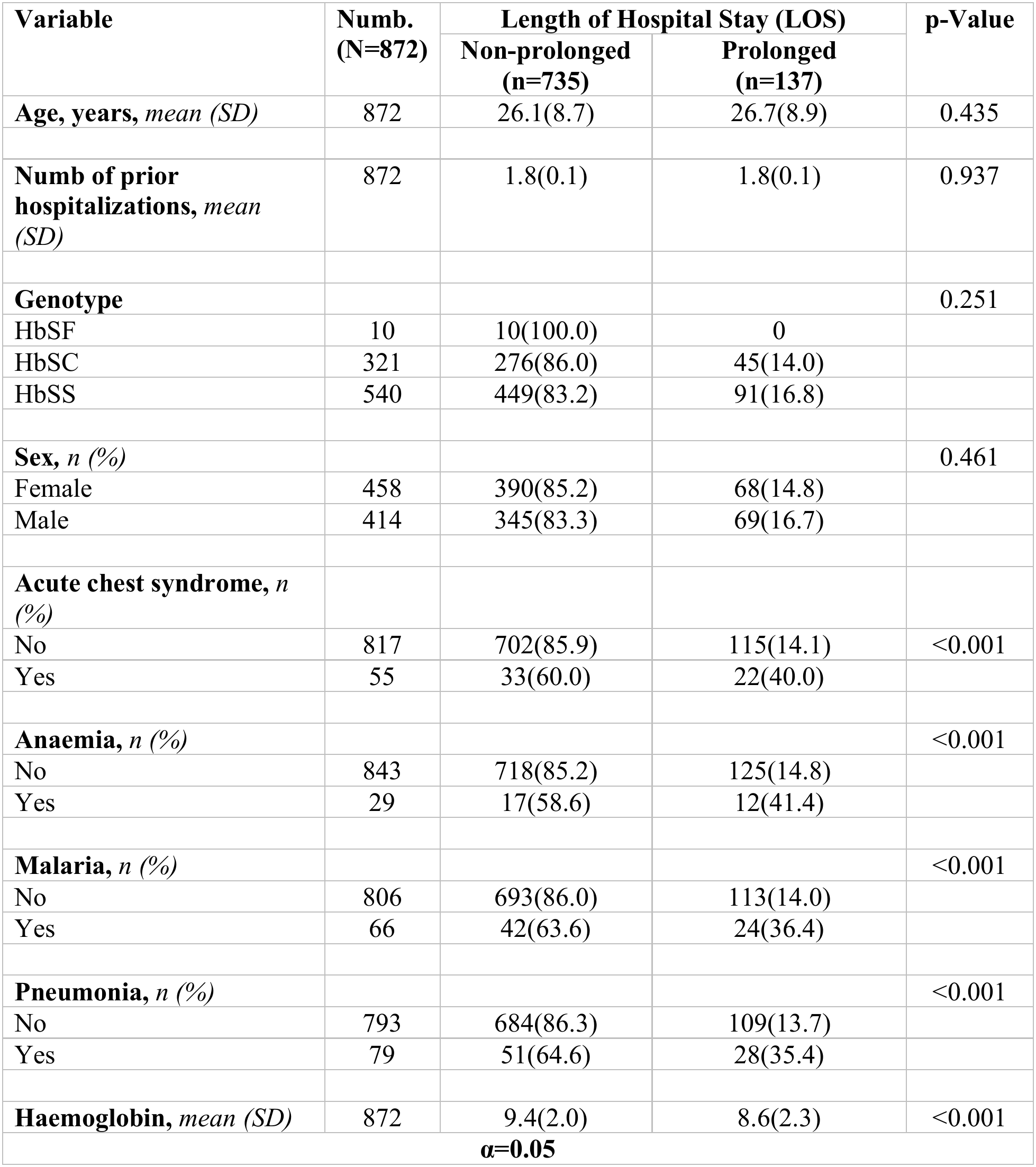
Baseline Sociodemographic and Clinical Characteristics of SCD inpatients, Stratified by Length of Hospital Stay.

| Variable | Numb. (N=872) | Length of Hospital Stay (LOS) |  | p-Value |
|---|---|---|---|---|
|  |  | Non-prolonged (n=735) | Prolonged (n=137) |  |
| **Age, years,** *mean (SD)* | 872 | 26.1(8.7) | 26.7(8.9) | 0.435 |
| **Numb of prior hospitalizations,** *mean (SD)* | 872 | 1.8(0.1) | 1.8(0.1) | 0.937 |
| **Genotype** |  |  |  | 0.251 |
| HbSF | 10 | 10(100.0) | 0 |  |
| HbSC | 321 | 276(86.0) | 45(14.0) |  |
| HbSS | 540 | 449(83.2) | 91(16.8) |  |
| **Sex,** *n (%)* |  |  |  | 0.461 |
| Female | 458 | 390(85.2) | 68(14.8) |  |
| Male | 414 | 345(83.3) | 69(16.7) |  |
| **Acute chest syndrome,** *n (%)* |  |  |  |  |
| No | 817 | 702(85.9) | 115(14.1) | <0.001 |
| Yes | 55 | 33(60.0) | 22(40.0) |  |
| **Anaemia,** *n (%)* |  |  |  | <0.001 |
| No | 843 | 718(85.2) | 125(14.8) |  |
| Yes | 29 | 17(58.6) | 12(41.4) |  |
| **Malaria,** *n (%)* |  |  |  | <0.001 |
| No | 806 | 693(86.0) | 113(14.0) |  |
| Yes | 66 | 42(63.6) | 24(36.4) |  |
| **Pneumonia,** *n (%)* |  |  |  | <0.001 |
| No | 793 | 684(86.3) | 109(13.7) |  |
| Yes | 79 | 51(64.6) | 28(35.4) |  |
| **Haemoglobin,** *mean (SD)* | 872 | 9.4(2.0) | 8.6(2.3) | <0.001 |
| **α=0.05** |  |  |  |  |

### Multivariable Logistic Regression

The multivariable logistic regression model identified acute chest syndrome, anaemia, malaria, pneumonia, and haemoglobin level as factors independently associated with prolonged hospitalization [Table 2]. Patients with acute chest syndrome had more than four-fold higher odds of prolonged hospitalisation compared with those without acute chest syndrome (aOR 4.28, 95% CI: 2.31–7.94, p < 0.001). Similarly, patients diagnosed with malaria had nearly four-fold higher odds of prolonged hospitalisation compared with those without malaria (aOR 3.77, 95% CI: 2.11–6.74, p < 0.001).

**Table 2.** Unadjusted and adjusted associations with prolonged hospitalisation (multivariable logistic regression)

| **Characteristic** | **Crude Odds Ratio** |  |  | **Adjusted Odds Ratio** |  |  |
|---|---|---|---|---|---|---|
|  | **OR** | **95% CI** | **p-Value** | **OR** | **95% CI** | **p-Value** |
| **Acute chest syndrome** |  |  |  |  |  |  |
| No | Ref |  |  | Ref |  |  |
| Yes | 4.06 | 2.29–7.23 | <0.001 | 4.28 | 2.31–7.94 | <0.001 |
| **Anaemia** |  |  |  |  |  |  |
| No | Ref |  |  | Ref |  |  |
| Yes | 4.05 | 1.89–8.69 | <0.001 | 2.71 | 1.13–6.51 | 0.026 |
| **Malaria** |  |  |  |  |  |  |
| No | Ref |  |  | Ref |  |  |
| Yes | 3.50 | 2.04–6.01 | <0.001 | 3.77 | 2.11–6.74 | <0.001 |
| **Pneumonia** |  |  |  |  |  |  |
| No | Ref |  |  | Ref |  |  |
| Yes | 3.44 | 2.08–5.70 | <0.001 | 3.45 | 2.02–5.90 | <0.001 |
| **Haemoglobin** | 0.81 | 0.74–0.89 | <0.001 | 0.86 | 0.78–0.96 | 0.005 |
| **Age at admission** | 1.00 | 0.99–1.02 | 0.435 | 1.00 | 0.97–1.02 | 0.711 |

**CI= Confidence Interval**

### Machine Learning Model Comparison

Table 3 presents the summaries of the test-set performance of the five supervised ML algorithms. Logistic regression demonstrated the highest discrimination (AUC-ROC=0.692, 95% CI 0.584–0.795), while Random Forest achieved the highest AUC-PR (0.375, 95% CI 0.223–0.542). Logistic regression achieved the highest sensitivity among the three leading models (51.9%), F1-score (0.418), and balanced accuracy (0.677). KNN achieved the highest sensitivity (59.3%) but had lower specificity (71.6%) and PPV (27.6%). XGBoost achieved the highest specificity (87.2%) but the lowest AUC-ROC (0.627) and sensitivity (40.7%). Confidence intervals for AUC-ROC overlapped substantially across all five models, indicating no clear evidence of superior discrimination among the algorithms.

**Table 3:** Test-set performance of the five supervised machine learning algorithms.

| Model | AUC-ROC (95% CI) | AUC-PR (95% CI) | Sensitivity | Specificity | PPV | NPV | F1<br>(95% CI) | Balanced Accuracy (95% CI) | Brier Score |
|---|---|---|---|---|---|---|---|---|---|
| Logistic Regression | **0.692**<br>**(0.584-0.795**) | 0.353<br>(0.203-0.523) | 0.519 | 0.831 | 0.359 | 0.904 | **0.418**<br>**(0.259-0.571)** | **0.677**<br>**(0.579-0.776)** | 0.205 |
| Support Vector Machine | 0.691<br>(0.578-0.803) | 0.364<br>(0.212-0.537) | 0.444 | 0.831 | 0.324 | 0.891 | 0.374<br>(0.227-0.523) | 0.640<br>(0.543-0.741) | 0.210 |
| Random Forest | 0.688<br>(0.569-0.795) | **0.375**<br>**(0.223-0.542)** | 0.481 | 0.824 | 0.333 | 0.897 | 0.391<br>(0.233-0.538) | 0.655<br>(0.553-0.761) | 0.221 |
| K-nearest Neighbour | 0.654<br>(0.527-0.773) | 0.310<br>(0.165-0.457) | **0.593** | 0.716 | 0.276 | **0.906** | 0.374<br>(0.240-0.505) | 0.656<br>(0.552-0.750) | **0.201** |
| XGBoost | 0.627<br>(0.497-0.751) | 0.303<br>(0.169-0.473) | 0.407 | **0.872** | **0.367** | 0.890 | 0.380<br>(0.213-0.540) | 0.641<br>(0.545-0.742) | 0.222 |

### Confusion Matrices

Figure 2 presents the confusion matrices for all the five models evaluated at the default probability threshold of 0.5. Across models, a substantial proportion of patients with prolonged hospitalisation were incorrectly classified as having non-prolonged hospitalisation. For example, the random forest model correctly identified 13 of the 27 patients with prolonged hospitalisation in the test set, corresponding to a sensitivity of 48.1%.

**Figure 2.**
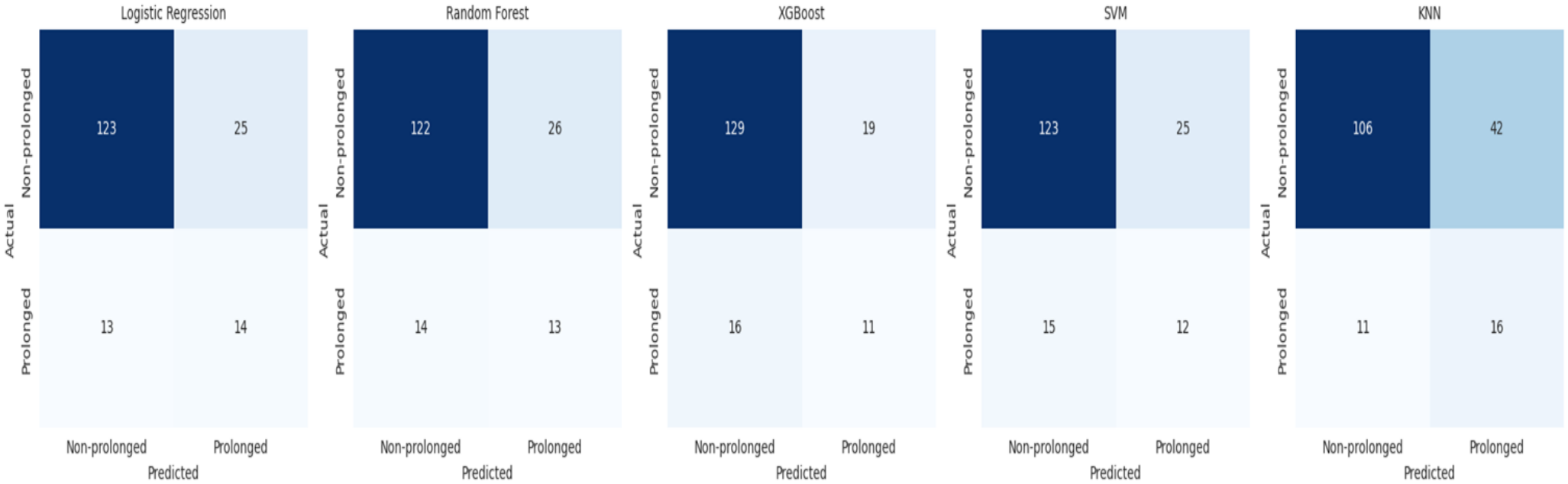
Confusion matrices for all five models on the held-out test set.

### Feature Importance

Figure 3 presents permutation feature importance for each algorithm. Pneumonia and haemoglobin were consistently the most influential predictors across most algorithms. On the other hand, Acute chest syndrome (x=0.018; SD=0.014) and anaemia (x=0.006; 0.017) contributed comparatively less to model discrimination in the ML models

**Figure 3.**
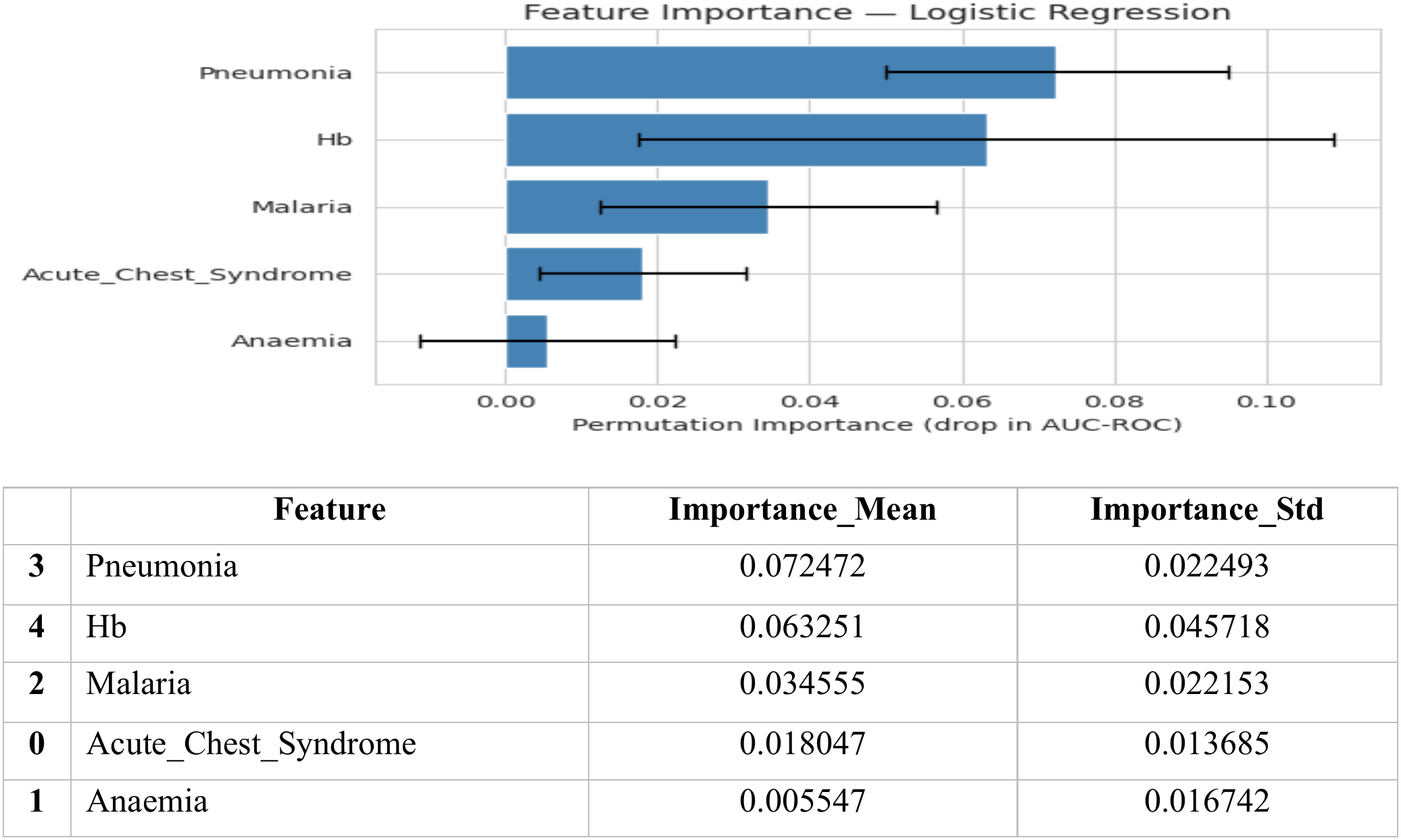
Permutation feature importance by algorithm.

## DISCUSSION

This study assessed factors associated with prolonged hospitalisation (>7 days) among patients with SCD and compared five supervised ML algorithms with conventional logistic regression for predicting this outcome. Prolonged hospitalisation occurred in 15.7% of admissions. Acute chest syndrome, malaria, pneumonia and anaemia were independently associated with higher odds of prolonged stay. A unit increase in haemoglobin, on the other hand, was associated with a 14% reduction in odds for prolonged hospitalization. The classification performance was modest across the models as the logistic regression had the highest AUC-ROC, F1-score and balanced accuracy, while Random Forest had the highest AUC-PR, KNN neighbor the highest sensitivity and XGBoost the highest specificity and PPV.

The prevalence of prolonged hospitalisation in our facility was 15.7% (95% CI: 13.4% to 18.2%), meaning approximately one in six admissions stayed beyond 7 days. This is comparable to the 16% reported at Komfo Anokye Teaching Hospital (KATH) in Ghana, although that study included patients aged ≥13 years(7). The similarity across two major Ghanaian hospitals suggests that prolonged admission is a consistent and substantial burden among patients with SCD, rather than a single-centre phenomenon. However, direct comparison should be cautious because disease severity, genotype distribution and availability of specialist differ across the two health facilities. The observed burden reinforces the need for effective interventions that prevents prolonged hospitalization among SCD patients; especially that prolonged hospitalization is linked with poor outcomes such as nosocomial infections, economic burden and caregiver-stress(5,6,10).

Acute chest syndrome (ACS) was significantly associated with increased odds of prolonged hospitalisation among patients with SCD. This finding is consistent with the study in KATH that reported higher odds of prolonged hospitalisation among SCD patients with ACS (7). ACS as a severe, acute lung injury, prolongs hospitalization because its management typically necessitates aggressive oxygen therapy, blood transfusions and intensive monitoring before a patient reaches clinical stability.

Patients with pneumonia were 3.5 times more likely to experience a prolonged hospital stay compared with patients without pneumonia. The association with pneumonia is clinically plausible because pulmonary infection can worsen hypoxemia and systemic inflammation, thereby promoting erythrocyte sickling and vaso-occlusive complications (11). Furthermore, treating pneumonia requires prolonged antimicrobial therapy, respiratory support and close clinical monitoring, all of which extend the time to recovery.

Having malaria increased the SCD patients’ odds for prolonged hospitalization by 3.8 times, compared with patients without malaria. This finding aligns with the broader clinical consensus that malaria acts as a severe pathological catalyst in individuals with homozygous sickle cell anaemia, triggering acute haemolytic crises, worsening baseline chronic anaemia, and precipitating vaso-occlusive events(12). Because malaria infection promotes systemic inflammation and microvascular sludging, patients frequently develop severe multisystem complications that require extended antimalarial therapy, intensive hydration, and frequent blood transfusions, thereby prolonging their hospital course.

Anaemia was a significant predictor of prolonged hospitalisation in our cohort, likely reflecting low haemoglobin as a marker of severe SCD rather than an isolated cause. Consistent with Ershler et al. (2023), anaemia may initiate a cascade of complications and greater healthcare utilisation(13). Each 1 g/dL increase in haemoglobin was associated with a 14% reduction in the odds of prolonged hospitalisation, suggesting haemoglobin is a useful indicator of disease burden and clinical stability. These findings support monitoring and optimising haemoglobin in hospitalised patients with SCD.

The traditional and machine-learning models demonstrated modest predictive performance with no clear differences in discrimination. The comparable performance of logistic regression may reflect predominantly linear predictor–outcome relationships and the small number of predictors under which more complex algorithms may offer limited incremental benefit(14). Under such condition, logistic regression provides a pragmatic and interpretable approach for risk stratification, while alternative algorithms may be preferable when sensitivity, specificity, or minority-class discrimination is prioritized.

### 4.1 Limitations and Strengths

There are limitations worth sharing. First, this was a retrospective, single-centre study, which may limit generalisability to other settings. Second, the sample size (n=872) is modest for ML model development, particularly for the minority class(n=137). Third, potentially relevant predictor like socioeconomic status was not available and may confound or mediate the associations observed.

## 5. Conclusion

The multivariate logistic regression indicates that infection-related complications (acute chest syndrome, malaria, pneumonia) and anaemia severity are key independent determinants of prolonged hospitalisation among SCD inpatients in this malaria-endemic cohort while higher haemoglobin was protective. Supervised ML algorithms offered modest, and not clearly superior, discrimination compared with conventional logistic regression at this sample size. These findings support infection prevention and haemoglobin optimisation as potential targets for reducing prolonged hospitalisation, and highlight the need for larger and externally validated studies before predictive models are considered for clinical use.

## Data Availability

De-identified data may be made available to qualified researchers upon reasonable request, subject to approval by the relevant institutional ethics committee and the University of Ghana Hospital and execution of the required data-sharing agreement.

## REFERENCES

1. Drahos J, Boateng-Kuffour A, Calvert M, Valentine A, Mason A, Li N, et al. Health-related quality of life and economic impacts in adults with sickle cell disease with recurrent vaso-occlusive crises: findings from a prospective longitudinal real-world survey. Qual Life Res [Internet]. 2025 Jul 17;34(7):2019–29.

2. WHO. Factsheet. 2025 [cited 2026 Mar 3]. p. 1–5 Sickle-cell disease Key facts. Available from: https://www.who.int/news-room/fact-sheets/detail/sickle-cell-disease

3. Thomson AM, McHugh TA, Oron AP, Teply C, Lonberg N, Vilchis Tella V, et al. Global, regional, and national prevalence and mortality burden of sickle cell disease, 2000–2021: a systematic analysis from the Global Burden of Disease Study 2021. Lancet Haematol

4. Awad A, Bader–El–Den M, McNicholas J. Patient length of stay and mortality prediction: A survey. Heal Serv Manag Res. 2017;30(2).

5. Rizwan M, Ikram A, Khan MQA, Baig ZF, Abid M, Bashir A. Outcomes of Healthcare Associated Infection in Intensive Care Units of A Tertiary Care Hospital. Pakistan Armed Forces Med J. 2025;75.

6. Pérez-Corraliza C, Castaño-Blanco MY, Romero-Perea JM, Muñoz-Bermejo L. Factors Associated with the Need for Support Among Informal Caregivers During Prolonged Hospitalization. Healthcare [Internet]. 2026 Aug 6;14(15):2420. Available from: https://www.mdpi.com/2227-9032/14/15/2420

7. Gyan KF, Gyabaah S, Ahmed EA, Osei L, Naabo MN, Owiredu MA, et al. Beyond Childhood: Adult and Adolescent Sickle Cell Disease and Outcomes in Northern Ghana. eJHaem. 2025;6(2).

8. Lee J. An insight on the use of multiple logistic regression analysis to estimate association between risk factor and disease occurrence. Int J Epidemiol. 1986;15(1).

9. Prasad S, Murphy SA, Morrow DA, Scirica BS, Sabatine MS, Berg DD, et al. Application of machine learning and deep learning approaches for prediction modeling with time-to-event outcomes in clinical epidemiology. Methods comparison and practical considerations for generalizability and interpretability. Ann Epidemiol. 2025;111.

10. Win KHH, Sule-saa S, Towfig MF, Kotei R, Nguyen KT, Hein PP, et al. The impact of viral pneumonia on hospital outcomes in sickle cell disease: A database review and analysis. Blood. 2025;146(Supplement 1):1196–1196.

11. Ahmed SG. The role of infection in the pathogenesis of vaso-occlusive crisis in patients with sickle cell disease. Mediterr J Hematol Infect Dis. 2011;3(1).

12. Uyoga S, Olupot-Olupot P, Connon R, Kiguli S, Opoka RO, Alaroker F, et al. Sickle cell anaemia and severe Plasmodium falciparum malaria: a secondary analysis of the Transfusion and Treatment of African Children Trial (TRACT). Lancet Child Adolesc Heal. 2022;6(9).

13. Ershler WB, De Castro LM, Pakbaz Z, Moynahan A, Weycker D, Delea TE, et al. Hemoglobin and End-Organ Damage in Individuals with Sickle Cell Disease. Curr Ther Res - Clin Exp. 2023;98.

14. de Groot LM, Twisk JWR, Kok AAL, Heymans MW. Incorporating longitudinal variability in prediction models: A comparison of machine learning and logistic regression in a cohort study with long follow-up. Ann Epidemiol. 2025;110.

